# Whole-Genome Promoter Profiling of Plasma Cell-Free DNA Exhibits Predictive Value for Preterm Birth

**DOI:** 10.1101/2022.09.20.22280143

**Authors:** Zhiwei Guo, Ke Wang, Xiang Huang, Kun Li, Guojun Ouyang, Xu Yang, Jiayu Tan, Haihong Shi, Liangping Luo, Xincai Zhang, Min Zhang, Bowei Han, Xiangming Zhai, Yingsong Wu, Fang Yang, Xuexi Yang, Jia Tang

## Abstract

Preterm birth (PTB) occurs in around 11% of all births worldwide, resulting in significant morbidity and mortality for both mothers and offspring. Identification of pregnancies at risk of preterm birth in early pregnancy may help improve intervention and reduce its incidence. However, there exist few methods for PTB prediction developed with large sample size, high throughput screening and validation in independent cohorts. Here, we established a large-scale, multi-center, and case-control study that included 2,590 pregnancies (2,072 full-term and 518 preterm pregnancies) from three independent hospitals to develop a preterm birth classifier. We implemented whole-genome sequencing on their plasma cfDNA and then their promoter profiling (read depth spanning from −1 KB to +1 KB around the transcriptional start site) was analyzed. Using three machine learning models and two feature selection algorithms, classifiers for predicting preterm delivery were developed. Among them, a classifier based on the support vector machine model and backward algorithm, named PTerm (**<u>P</u>**romoter profiling classifier for pre**<u>term</u>** prediction), exhibited the largest AUC value of 0.878 (0.852–0.904) following LOOCV cross-validation. More importantly, PTerm exhibited good performance in three independent validation cohorts and achieved an overall AUC of 0.849 (0.831–0.866). Taken together, PTerm could be based on current noninvasive prenatal test (NIPT) data without changing its procedure or adding detection cost, which can be easily adapted for preclinical tests.

## Introduction

Preterm birth (PTB) is a common complication of pregnancy, which was observed in around 11.1% of newborns worldwide ^1^. In addition, PTB is the main determinant of infant morbidity and mortality, which is responsible for approximately 35% of pregnancy-related deaths and can also lead to adverse maternal and fetal outcomes with increased long-term risks of complications, such as motor, cognitive and behavioral disorders ^2^. Importantly, the development of interventions to prevent premature delivery requires early identification of pregnant women at risk before preterm birth occurs. Since the maternal circulatory system carries both maternal and fetal information, multivariate screening methods based on maternal blood omics data, such as metabolite and cell-free RNA (cfRNA) ^3, 4, 5^, have recently been proposed. However, to date there is still a lack of reliable biomarkers for pregnancy complications, making the identification of PTB biomarkers a critical priority.

Plasma cell-free DNA (cfDNA) has been widely used in many clinical applications demonstrating its stability and applicability in clinics ^6, 7^. In pregnancy, the proportion of cell-free fetal DNA (cffDNA) in maternal cfDNA is an important feature of cfDNA. Pregnancies who have an elevation of cffDNA are at increased risks for preterm birth delivery ^8^. But changes in cffDNA levels have also been observed in many other pregnancy complications, such as preeclampsia ^9^. Taken together, these studies suggest that cfDNA has significant potential as a non-invasive biomarker for diverse diseases. However, it is necessary to identify novel disease-specific cfDNA characteristics before applying them to the prediction of preterm birth in early pregnancy.

During pregnancy, plasma cfDNA is primarily derived from placental trophoblasts and hematopoietic cells and is released following their apoptosis during the enzymatic processing of the chromatin. The DNA bound to the nucleosome is retained, while the exposed DNA between nucleosomes is digested ^10, 11^. Thus, the resulting cfDNA comprises a nucleosome footprint carrying information about its tissues of origin ^10, 12^. For example, analysis of maternal plasma cfDNA and expression profiles of both the placenta and whole blood revealed that the promoter regions of active genes exhibited depleted read coverage in the cfDNA implying that the nucleosome was less tightly bound within the promoter regions along with higher gene expression level ^13^. In addition, preterm birth is a common complication resulting from placental dysfunction and changes in the maternal immune system ^8^. Therefore, we hypothesized that the distribution patterns of plasma cfDNA fragments may carry information regarding the source tissues of origin, particularly placental trophoblasts and maternal hematopoietic cells and that global profiling of cfDNA fragments in promoter regions can be applied to identify predictive biomarkers for preterm birth (Figure 1).

**Figure 1.**
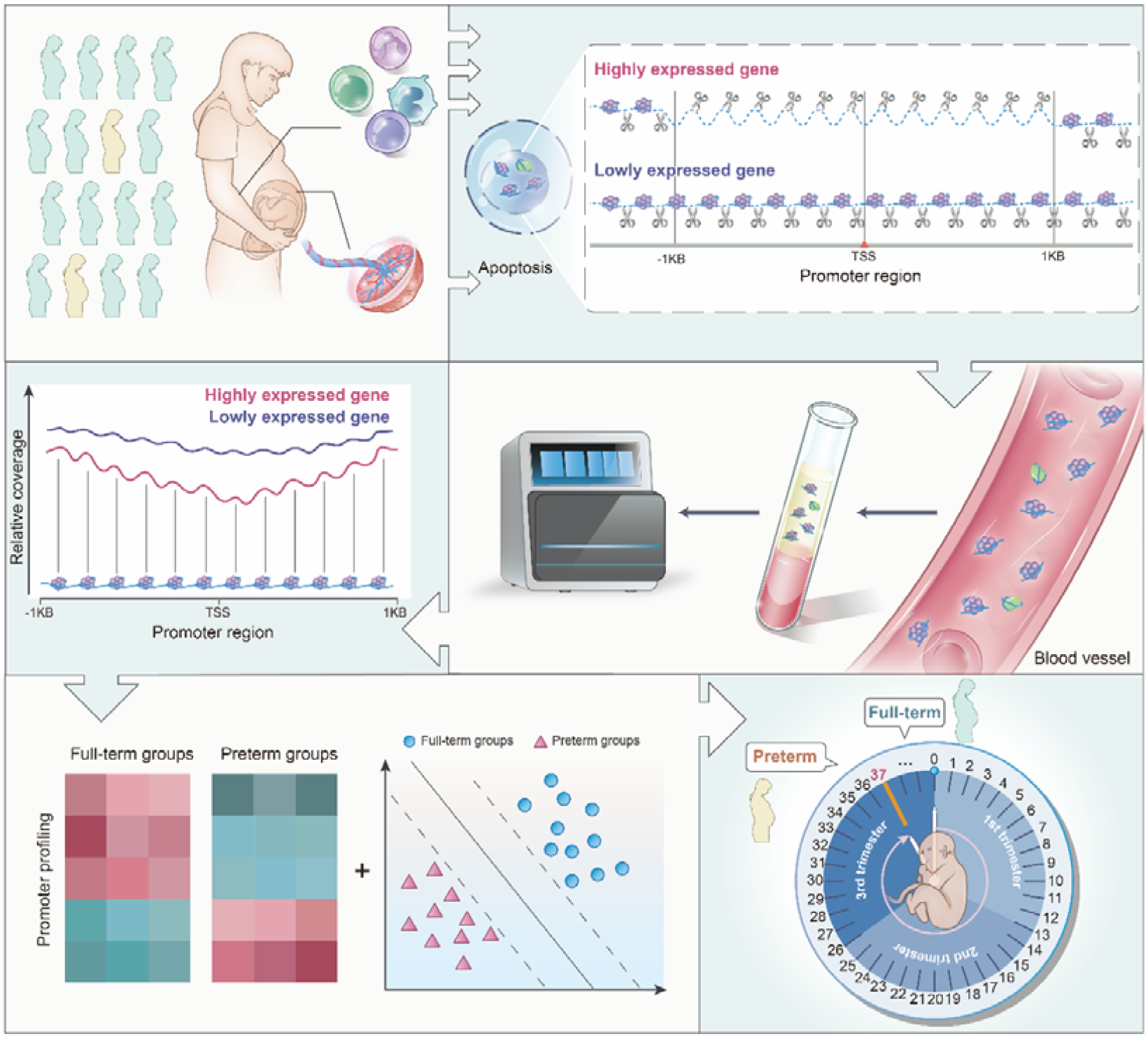
Schematic overview describing the prediction of preterm pregnancies using promoter profiling of plasma cfDNA. During pregnancy, the plasma cell-free DNA (cfDNA) is mainly derived from placental trophoblasts and maternal hematopoietic cells, which is released by their apoptotic cells. Exposed DNA not bound to a nucleosome is digested, whereas nucleosome-bound DNA escapes digestion and enters into the maternal circulation. By implementing whole-genome sequencing, we found that the read coverages at pTSS regions (−1KB to +1KB around the transcription start site [TSS]) could reflect the gene expression patterns of its tissues of origins. Since premature delivery is closely associated with dysfunction and changes in the placenta and maternal immune system, we proposed that cfDNA coverages at pTSS regions could be used to predict the occurrence of preterm birth at early gestational age. We tested this hypothesis using high-throughput whole-genome sequencing of plasma cfDNA from 2,590 preterm and full-term pregnancies across three independent hospitals. By comparing their promoter profiling, we found that the promoter profiling was different between preterm and full-term pregnancies. Then we used the genes with differential promoter coverage and three different machine learning models to develop predictive classifiers for PTB. To show greater differences, all nucleosomes in the promoter regions of highly expressed genes are depleted. The nucleosome-depleted region is usually found within the nucleosome upstream of the TSS.

In this study, we carried out a large-scale, retrospective study to develop classifiers for predicting preterm birth using whole-genome sequencing of plasma cfDNA from 2,590 pregnancies across three independent hospitals. Specific promoter profiling was found for preterm and full-term pregnancies. We applied three predictive models and two feature selection algorithms to develop classifiers that could predict the occurrence of preterm birth. Among these classifiers, a classifier that relied on the SVM model and backward algorithm, named PTerm (**<u>P</u>**romoter profiling classifier for pre**<u>term</u>** prediction), performed well as a predictor of PTB and exhibited an overall AUC value of 0.849 (0.831–0.866) among all cohorts. Our findings suggest that cfDNA coverage across certain promoter regions detected at early gestational age may be helpful in the development of simple and precise methods for the prediction of placenta-origin pregnancy complications.

## Results

### cfDNA carries information about its origin in pregnant women

Previous studies have reported that cfDNA carries information regarding its tissues of origin ^10, 11, 12, 13^, making it an ideal choice for evaluations around preterm birth. Thus, we designed these experiments to characterize the cfDNA profiles of pregnancies resulting in preterm and full-term births. To this end, we collected whole-genome sequencing data from 20 preterm and 20 full-term pregnancies (Supplemental Table S4). We also collected the RNA expression profiles of both placenta and whole blood from preterm pregnancies (GSE73685).

We first compared the read coverage at the pTSS between the 500 highest and 500 lowest expressed genes in the placenta, and found that the 500 most highly expressed genes showed reduced depth at the pTSS regions compared with the 500 least expressed genes (Figure 2a,b; *P*-value < 2.2e-16, Wilcoxon rank-sum test). In addition, the housekeeping genes with highly expressed levels exhibited lower read depth, whereas the unexpressed genes with lowly expressed levels exhibited higher read depth (Figure 2c,d; *P*-value < 2.2e-16, Wilcoxon rank-sum test). Similar patterns were also observed in maternal whole blood data (Supplemental Figure 1). Therefore, we confirmed that the coverages of plasma cfDNA at the pTSS regions were closely correlated with the expression profiles of its original tissues, suggesting that promoter profiling could reflect the expression status of its original tissues. Next, we focused on the cfDNA profiles of placenta-specific genes, which were closely related to placental functions. The results revealed that placenta-specific genes were characterized by reduced coverages at the pTSS regions in preterm birth pregnancies when compared to full-term pregnancies (Figure 1e; *P*-value < 2.2e-16, Wilcoxon rank-sum test), implying that there may be broad differences in the promoter profiling of these two patient groups.

**Figure 2.**
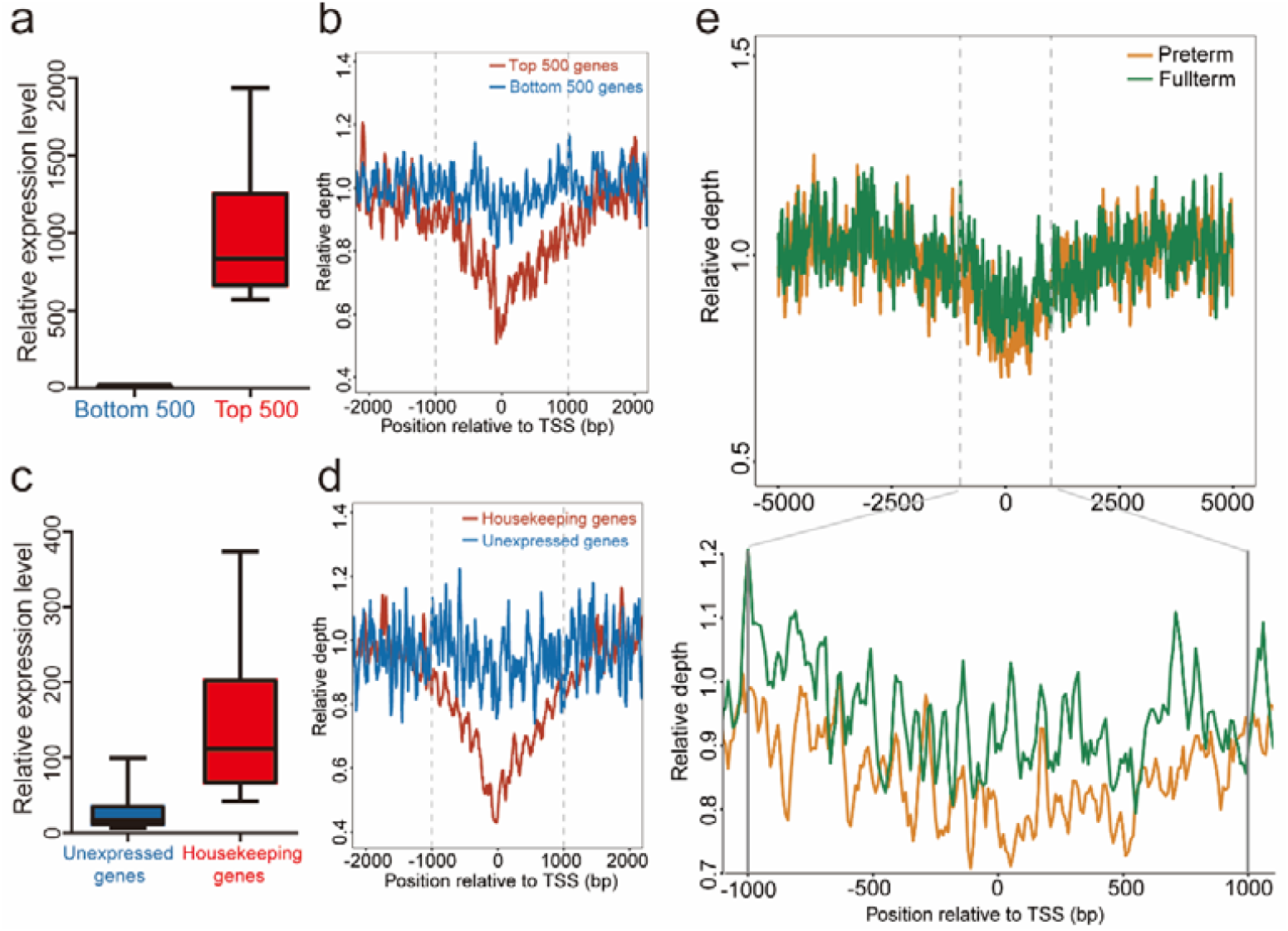
cfDNA profiles of the promoter regions reflect nucleosome positioning in pregnant women. **(a)** Average expression of the 500 most- (Top500, red) and least-expressed genes (Bottom500, blue) in the placenta of preterm birth pregnancies. **(b)** Read depth of whole-genome sequencing across pTSS regions (−1 KB to 1 KB around the TSS) from the 500 most- (Top500, red line) and least-expressed (Bottom500, blue line) genes. The read depth of the Top500 genes was lower than that of the Bottom500 genes (*P*-value < 2.2e-16, Wilcoxon rank-sum test). **(c)** Average expression levels of the housekeeping (red) and unexpressed (blue) genes in the placenta. **(d)** Read depth of whole-genome sequencing at the pTSS region (−1KB to +1KB around TSS) of the housekeeping genes (red line) was lower than that of unexpressed genes (blue line) in the placenta (*P*-value < 2.2e-16, Wilcoxon rank-sum test). **(e)** Sequencing read depth of placenta-specific genes was shown to be more depleted in the preterm (yellow line) pregnancies than full-term (green line) pregnancies (*P*-value < 2.2e-16, Wilcoxon rank-sum test). Placenta-specific, unexpressed, top500, and bottom500 genes are shown in Supplemental Table S1-S3. The pTSS region (−1KB to +1KB around the TSS) is denoted by the grey dashed lines. PTB = preterm birth, TSS = transcriptional start site, cfDNA = cell-free DNA.

### Promoter profiling of cfDNA reveals PTB-associated patterns

We then investigated whether cfDNA promoter profiling of preterm and full-term pregnancies demonstrated any deviations in the pattern. By comparing their cfDNA promoter profiling, we identified 277 genes with differential coverages at pTSS (Figure 3a; |Log2 fold change| > 1 and FDR < 0.05). These genes included 146 genes with increased coverage and 131 genes with decreased coverage (Supplemental Table S5). Next, we used PCA on these genes and found that the samples from the same group had similar promoter profiling (Figure 3b). More importantly, the application of unsupervised clustering analysis produced the heatmap that revealed distinct differences in promoter coverage for preterm and full-term pregnancies (Figure 3c).

**Figure 3.**
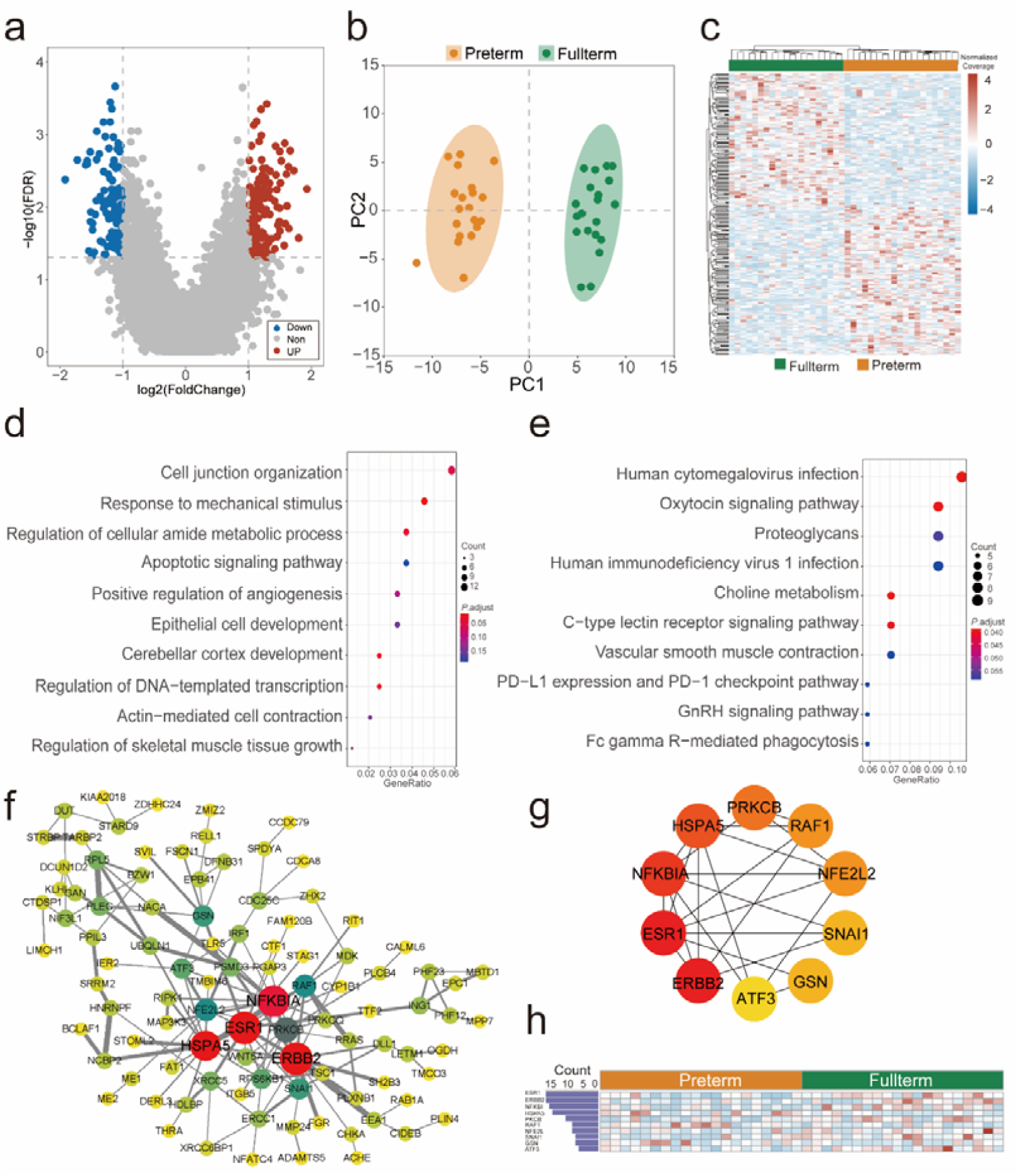
Differences in pTSS profiles of preterm and full-term pregnancies. **(a)** Volcano plots describing the gene transcripts with differential read coverages within the pTSS regions between 20 preterm birth (PTB) and 20 full-term pregnant women. A total of 277 transcripts with differential read coverages at pTSS regions were identified (|log_2_ fold change| ≥ 1 and false discovery rate [FDR] ≤ 0.05). The red, blue and grey dots indicate transcripts with increased, decreased, and non-differential coverage, respectively. The X- and Y-axes represent the log fold change and *P*-value calculated by the Wilcoxon rank-sum test, respectively. The raw *P-*value was adjusted to the false discovery rate (FDR) using the Benjamini-Hochberg procedure. **(b)** Principal Component Analysis (PCA) of these genes. **(c)** Heat map describing the z-scores of the genes with differential read coverages at pTSS as generated by the pheatmap package (ver. 1.0.2) when applied using the complete-linkage clustering algorithm. **(d)** Gene Ontology enrichment of transcripts with differential coverage between PTB and preterm birth groups using Metascape (ver. 20220101). **(e)** KEGG pathway enrichment of transcripts with differential coverage between PTB and term groups using clusterProfiler (ver. 3.18.1). **(f)** Gene correlation network for transcripts with differential coverage between PTB and term groups, with gene correlation evaluated using the String database (ver. 11.5) and network visualization by Cytoscape (ver. 3.8). Here, we merely showed the major correlation network. **(g)** Correlation network for the hub genes, with each hub’s degree of significance determined using cytohubba (ver. 0.1). **(h)** Degrees and heatmap of hub genes interconnection within the correlation network.

GO and KEGG enrichment analyses were then used to annotate the functions of the genes with differential coverages at pTSS. The results of GO enrichment showed that the GO terms associated with cell junction organization, response to mechanic stimulus, apoptosis, and development were closely related to embryonic development and premature delivery (Figure 3d). Taking the apoptotic signal pathway as an example, previous studies have shown that the apoptosis of fetal membranes could plausibly contribute to the risk of PTB ^14^. In addition, the results of KEGG enrichment analysis showed that a large proportion of the enriched pathways were closely associated with embryonic development and preterm birth (Figure 3e). As one example, premature activation of oxytocin secretion often results in preterm labor and oxytocin receptor antagonists could inhibit preterm birth ^15^. These results may suggest that the genes with differential coverage at pTSS may be a clinical indicator for PTB.

Finally, we tried to find the potential key genes associated with PTB using a gene correlation network. The analysis of gene functional connections allowed us to evaluate the degree of gene influence and importance (Figure 3f), which may help us identify the essential genes in the occurrence and progression of PTB. Our evaluations identified the top 10 hub genes according to their degree values. These genes included *ERBB2, ESR1, NFKBIA, HSPA5, PRKCB, RAF1, NFE2LE, SNAI1, GSN*, and *ATF3* (Figure 3h), which were then revealed to be associated with preterm birth, embryonic development and pregnancy (Supplemental Table S6) throughout the literature. Take *ESR1* as an example, its gene polymorphism is associated with premature delivery, with its DNA methylation patterns also showing distinct differences between preterm and full-term pregnancies ^16^. Furthermore, *ESR1* could regulate the *WNT4* to exert their effects on preterm birth ^17^. In particular, the expression of *ATF3* is significantly decreased in preterm placentas and *ATF3* is the regulator of soluble fms-like tyrosine kinase 1 (sFlt-1) and soluble Endoglin (sEng), which are important makers of premature delivery and preeclampsia ^18^.

### Promoter profiling of plasma cfDNA can predict preterm birth

To further validate the potential of cfDNA promoter profiling in predicting preterm birth, we established a large-scale, multi-center, and case-control study, which included 2,590 pregnant women, including 518 preterm and 2,072 full-term pregnancies, from three independent hospitals (Figure 4). Our training stage focused on the 277 gene transcripts with differential coverage at pTSS identified in the discovery stage. We then used three predictive models (SVM, LDA and LR) and two feature selection algorithms (backward and lasso algorithms) to develop the optimal predictive classifier. We found that the performance of the optimal classifiers for each model based on the backward feature selection algorithm was higher than those of classifiers with the lasso algorithm (Figure 5a–d and Supplemental Table 8; all *P*-value < 0.05, DeLong’s test). More importantly, we found that a classifier that relied on the SVM model and backward algorithm, named PTerm, performed well as the best predictor of PTB (accuracy = 87.3%, sensitivity = 88.5%, and specificity = 87.0%). PTerm exhibited the largest AUC value after LOOCV cross-validation (0.878 [0.852–0.904]) and its AUC value was higher than those of the optimal classifiers produced using the LDA and LR models (Figure 5a,b; all *P*-values < 0.05, DeLong’s test).

**Figure 4.**
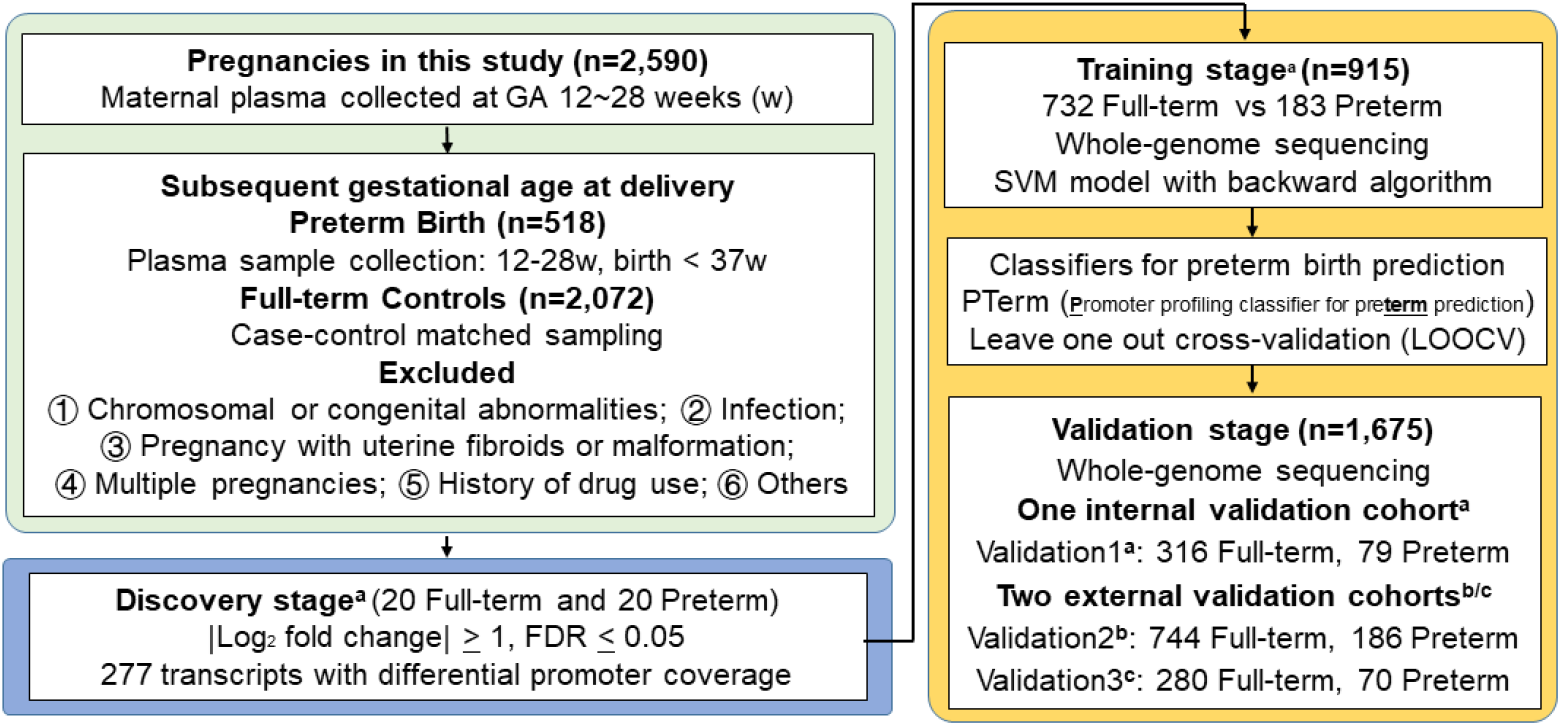
Pipeline for preterm birth classifier construction. In this study, 2,590 plasma cfDNA samples (518 preterm birth and 2,072 full-term pregnancies) were collected from three independent hospitals, including Jiangmen maternal & child healthcare hospital (JM)^a^, Foshan maternal & child healthcare hospital (FS)^b^, and Nanfang Hospital of Southern Medical University (NFY)^c^. These samples were collected at 12-28 weeks of gestation. According to their subsequent delivery time, the pregnant women were categorized as preterm or full-term groups. We then used the whole-genome sequencing data to develop classifiers for predicting PTB via three-step processes, including discovery, training, and validation. In the discovery stage, we identified 277 transcripts with differential coverage at pTSS regions (−1 KB to +1 KB around TSS) between these two groups. In the training stage, we applied non-linear support vector machine (SVM), linear discriminant analysis (LDA), and logistic regression (LR) models augmented with backward and lasso feature selection algorithms to develop a set of predictive classifiers. The performance of these classifiers was assessed using leave-one-out cross-validation method (LOOCV). The predictive classifier, denoted by PTerm, achieved the largest AUC was identified and its performance was further validated in the three validation cohorts, including one internal cohort (validation1, JM: n = 395) and two external cohorts (validation2 derived from FS: n = 930; Validation3 derived from NFY: n = 350). Additional details about participant definition and classifier construction are given in the method section and supplemental materials. PTB = preterm birth. GA = gestational age.

**Figure 5.**
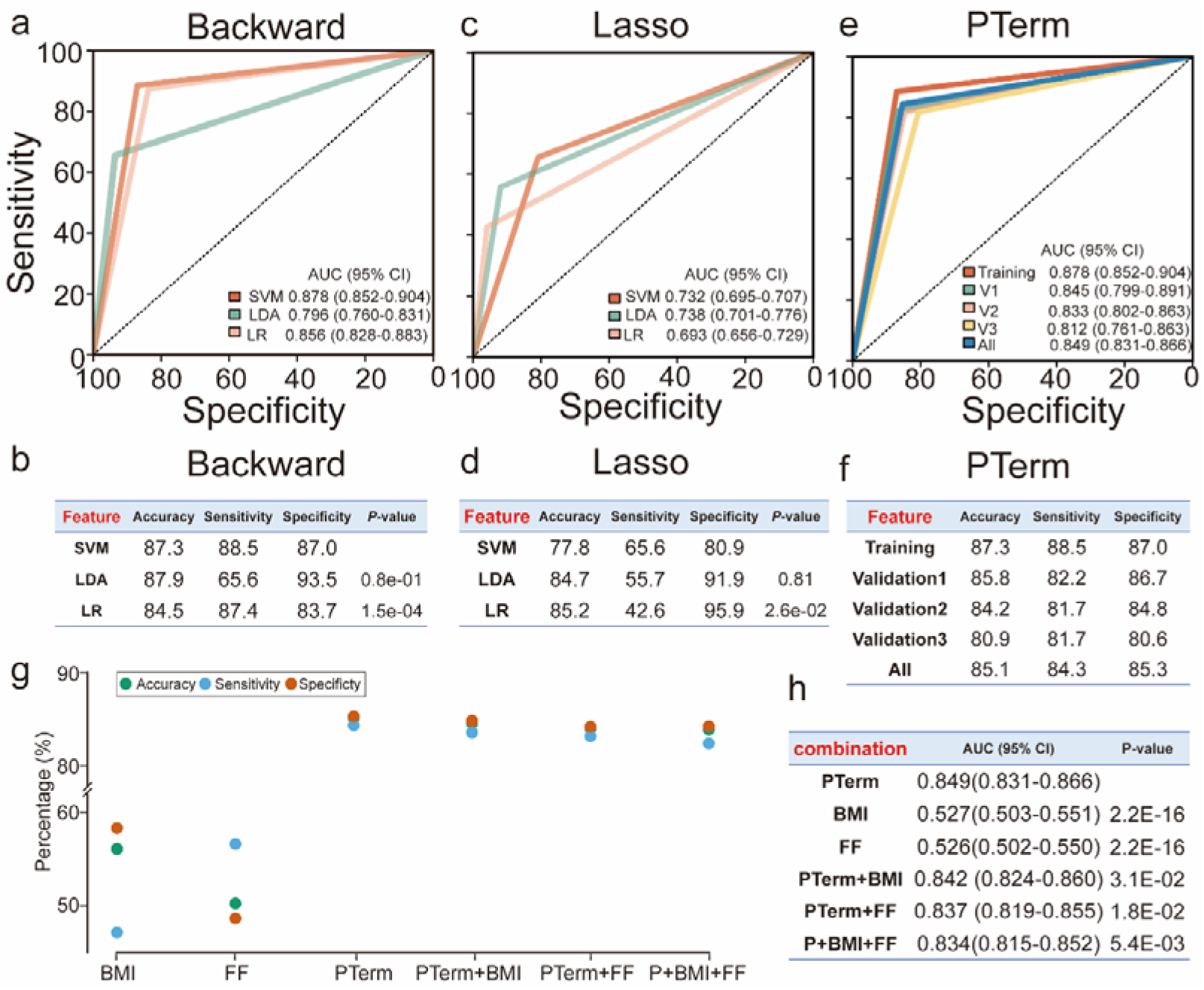
Performance of the classifiers in predicting preterm birth. **(a)** Receiver operating characteristic (ROC) curves for each of the predictive classifiers using backward feature selection algorithm. **(b)** Performance of each of the predictive classifiers using backward algorithm. **(c)** ROC curves for the predictive classifiers using the lasso feature selection algorithm. **(d)** Performance of the classifiers with lasso feature selection algorithm. **(e)** ROC curves of the optimal classifier, PTerm. **(f)** Performance of PTerm across each cohort. **(g)** Performance of different combination. **(h)** AUCs of different combination.

We then evaluated the performance of PTerm across three validation cohorts, including one internal and two external validation cohorts. Consistent with the results of the training cohort, PTerm exhibited solid predictive capacity in all three cohorts. The AUC for the internal validation1 cohort was 0.845 (0.799–0.891), and the AUCs for external validation2 and validation3 cohorts were 0.833 (0.802–0.863) and 0.812 (0.761–0.863), respectively (Figure 5e,f). In addition, PTerm produced an AUC of 0.849 (0.831–0.866) across all cohorts when discriminating between preterm and full-term pregnancies with a sensitivity of 84.4% and specificity of 85.3% (Figure 5e,f).

### PTerm combined with clinical features

Previous studies have reported that certain clinical features could be applied to predict preterm birth, such as fetal fraction (FF) and BMI before pregnancy (BMI). In our data, we found that the AUCs of BMI (0.527 [0.503–0.551]) and FF (0.526 [0.502–0.550]) were significantly lower than that of PTerm (Figure 5g,h; all *P*-value < 0.05, DeLong’s test). To attempt to improve the performance of our classifier, we further combined BMI and FF with PTerm. The AUCs of the combined classifiers (PTerm+BMI, PTerm+FF and PTerm+BMI+FF) were 0.842 (0.824–0.860), 0.837 (0.819–0.855) and 0.834 (0.815–0.852), which were also significantly lower than that of PTerm (Figure 5g,h; all *P*-value < 0.05, DeLong’s test).

## Discussion

In this study, we described the application of the promoter profiling of plasma cfDNA to predict premature delivery. We found that promoter profiling of cfDNA could reflect the expression status of its tissues of origins and broad changes in promoter profiling were observed between preterm and full-term pregnancies. Given this, we hypothesized that the differential read-depth patterns of cfDNA at promoters should supply sufficient information regarding placenta-origin diseases way before any clinical symptoms would appear (Figure 1). Thus, we developed a series of predictive classifiers using data from large-scale multi-center cohorts (n=2,590), which included pregnancies across three independent centers (Figure 4). These classifiers were developed using three different machine learning models and two different feature selection methods to ensure optimal performance. This development produced a robust prediction classifier, PTerm, which was shown to be able to predict PTB with an overall AUC of 0.849 (0.831–0.866). These findings highlight the potential values of promoter profiling of cfDNA as a non-invasive assessment for predicting preterm delivery at early gestational age.

Recent studies have made pioneering attempts to use maternal blood omics data (cfDNA, cfRNA and metabolite) to predict future complications in pregnancy, such as preterm birth and preeclampsia ^3, 4, 5, 13, 19, 20^. A biomarker study requires a large sample size, high-throughput screening, and independent cohort validation. So far, few studies have recruited more than 2,500 samples with high-throughput screening, and performed validation in multiple independent cohorts. In this study, 2,590 whole-genome sequencing of plasma cfDNA derived from 518 preterm and 2,072 full-term pregnancies were collected from three independent hospitals to train and validate the classifiers for predicting preterm birth. In addition, useful biomarkers for disease prediction need to be stable, non-invasive, and low-cost. cfDNA meets these needs and the detection of cfDNA (noninvasive prenatal test, NIPT) has been widely used for fetal trisomy detection worldwide. In 2018, 10 million NIPT tests were performed in more than 60 countries ^21^. Since PTerm could be based on current NIPT data without changing its procedure or adding detection cost, it can be easily adapted for preclinical tests. However, previous studies have revealed that the risk factors of premature delivery among different ethnic backgrounds tend to vary. More samples from other ethnicities are needed to test whether PTerm is suitable for other ethnic backgrounds.

There exist 83 genes in the PTerm classifier and these genes were closely associated with development, pregnancy and premature delivery (Supplemental Table S7). Firstly, these 83 genes were selected from the genes with differential coverages, of which GO and KEGG enrichment results showed that a substantial portion of GO terms and pathways were closely related to preterm birth, including apoptotic and oxytocin signaling pathways (Figure 3d,e). In addition, 4 of the 10 hub genes (*ERBB2, NFKBIA, RAF1* and *GSN*) were retained in the classifiers and their changes in expression levels or DNA polymorphism were closely related to preterm birth (Supplemental Table S6). Take *NFKBIA* as an example, the degradation of IκBα could activate NF-κB resulting in production of proinflammatory IL-6 and inflammation is closely associated with preterm birth ^22^. More importantly, research revealed that a substantial proportion of the genes in the PTerm were closely associated with development, pregnancy and premature delivery (Supplemental Table S9). These results may indicate that cfDNA promoter profiling can not only be used to predict preterm birth, but also help to determine potential therapeutic targets.

In summary, our data suggest that promoter-profiling based classifier (PTerm) could provide valuable PTB predictions in early pregnancy. Our method is also easily applicable to routine NIPT data and does not require any additional tests or detection costs making it feasible in clinical practice. Given this, we believe that our method provides a critical stepping stone toward the development of a non-invasive diagnostic for the early prediction of pregnancy complications.

## Methods

### Participant characteristics

In total, we collected 2,590 plasma samples from preterm and full-term pregnancies. These plasma samples were collected at 12–28 weeks of gestation from three independent hospitals across China, including Jiangmen maternal & child healthcare hospital (JM), Foshan maternal & child healthcare hospital (FS), and Nanfang hospital of southern medical university (NFY). These samples were then retrospectively assigned to a birth outcome group based on their subsequent delivery time with PTB defined as birth < 37 weeks of gestation. More details about the definition and selection of preterm birth and full-term controls are shown in the Supplemental materials. Of the 2,590 participants, 518 women experienced a preterm delivery while the remaining 2,072 women delivered at full-term (Table 1). The participants from JM were collected between Jan 2017 and Dec 2020. The participants enrolled at FS, were recruited between Dec 2018 and Dec 2020. The participants from NFY enrolled between May 2016 and May 2020.

**Table 1.** Clinical characteristics of pregnancies in four cohorts.

|  | Training<br>cohort<br>(n=915) | Validation<br>cohort1<br>(n=395) | Validation<br>cohort2<br>(n=930) | Validation<br>cohort3<br>(n=350) | *P*-value |
| --- | --- | --- | --- | --- | --- |
| Full-term pregnancies |  |  |  |  |  |
| n | 732 | 316 | 744 | 280 |  |
| Gestational age at<br>sampling (weeks) | 15.1±3.2 | 15.3±3.3 | 15.5±3.5 | 15.5±3.7 | 0.353 |
| Gestational age of<br>birth (weeks) | 39.7±0.8 | 39.7±0.8 | 39.6±0.8 | 39.6±0.8 | 0.264 |
| Maternal age (years) | 29.8±4.7 | 30.3±4.6 | 30.5±4.1 | 30.5±4.6 | 0.056 |
| BMI (kg/m<sup>2</sup>) | 20.8±2.8 | 20.6±2.9 | 20.6±1.1 | 20.6±2.7 | 0.479 |
| Birth length (cm) | 49.1±1.5 | 49.1±1.4 | 49.2±0.6 | 49.1±1.4 | 0.491 |
| Birth weight (kg) | 3.2±0.3 | 3.2±0.3 | 3.2±0.1 | 3.2±0.3 | 0.100 |
| Previous Birth, No. (%) |  |  |  |  | 0.099 |
| 0 | 312 (42.6%) | 147 (46.5%) | 329 (44.2%) | 128 (45.7%) |  |
| 1 | 392 (53.6%) | 161 (50.9%) | 370 (49.7%) | 143 (51.1%) |  |
| ≥2 | 28 (3.8%) | 8 (2.6%) | 45 (6.1%) | 9 (3.2%) |  |
| Preterm Pregnancies |  |  |  |  |  |
| n | 183 | 79 | 186 | 70 |  |
| Gestational age at<br>sampling (weeks) | 15.1±3.2 | 15.3±3.3 | 15.5±3.5 | 15.5±3.5 | 0.803 |
| Gestational age of<br>birth (weeks) | 35.3±1.8 | 35.2±1.7 | 34.7±2.6 | 34.7±2.6 | 0.066 |
| Maternal age (years) | 30.7±5.1 | 30.1±4.8 | 30.5±4.8 | 30.5±4.8 | 0.797 |
| BMI (kg/m<sup>2</sup>) | 21.2±3.1 | 21.1±3.0 | 20.6±1.2 | 20.6±1.2 | 0.619 |
| Birth length (cm) | 46±2.7 | 45.9±3.0 | 46.1±1.6 | 46.1±1.6 | 0.345 |
| Birth weight (kg) | 2.46±0.51 | 2.42±0.47 | 2.47±0.37 | 2.47±0.37 | 0.768 |
| Previous Birth, No. (%) |  |  |  |  | 0.069 |
| 0 | 82 (44.8%) | 32 (40.5%) | 98 (52.7%) | 29 (41.4%) |  |
| 1 | 94 (51.4%) | 43 (54.4%) | 73 (39.2%) | 35 (50.0%) |  |
| ≥2 | 7 (3.2%) | 4 (5.1%) | 17 (9.1%) | 6 (8.6%) |  |

Data are mean ± standard deviation. BMI = pre-pregnancy body mass index. Wilcoxon rank-sum
test was used for the comparison of continuous variables. Pearson $\chi^2$ test and Fisher exact test (\*) were used for the comparison of categorical variables.

All participants were singleton pregnancies and pregnancies were excluded: (1) chromosomal or congenital abnormalities; (2) infection; (3) pregnancies with uterine fibroids or uterine malformation; (4) history of heparin, aspirin, or other drug use. Gestational age was determined according to the last menstrual period. The institutional ethics committees of all hospitals approved this retrospective analysis, and the requirement ethics committees abandoned the requirement of informed consent.

### DNA-Seq processing and promoter profiling analysis

The procedure of sample collection, cfDNA isolation and DNA sequencing are in the Supplemental Materials. We estimated the fetal fraction using the proportion of all sequencing reads from the Y chromosome or the seqFF method. Gene information was obtained from the RefSeq of the University of California Santa Cruz (UCSC) Genome Browser Database ^23^. For each transcript, the promoter region, spanning from −1 KB to +1 KB around the transcriptional start site, was defined as pTSS regions. The pTSS regions found to overlap with the Duke blacklist region were removed (http://hgdownload.cse.ucsc.edu/goldenpath/hg19/encodeDCC/wgEncodeMapability/). After sequencing, the raw reads were aligned to the human reference genome, hg19, using bwa-mem (ver. 0.7.4). The polymerase chain reaction (PCR) duplicates were removed using the rmdup function of SAMtools (ver. 1.2). The GC-bias correction was implemented using deeptools (ver. 3.5.0) with its default settings. The read coverage for each pTSS region was extracted using bedtools (ver. 2.17.0). We then normalized the read coverage data using the following formula.

### Gene expression profile analysis and gene information acquisition

Placenta and whole blood expression profiles for preterm pregnancies (GSE73685) were downloaded from the Gene Expression Omnibus (GEO) database ^24^ and then normalized using GEOquery (ver. 3.3.1). The 500 most highly expressed genes and least expressed genes in the placenta and whole blood were then identified by analyzing their expression profiles (Supplemental Table S1). Placenta- and blood-specific genes were annotated using PaGenBase (Supplemental Table S2). Housekeeping and unexpressed genes ^11^ were obtained from the supplemental materials of previous studies (Supplemental Table S3).

### Genes with significant differential promoter coverages

At the discovery stage, we selected 20 PTB cases and 20 full-term pregnancies (Supplemental Table S4) and then completed whole-genome sequencing of their cfDNA. After data processing and normalization, pTSS coverages between preterm and full-term samples were compared to calculate the *P*-value using Wilcoxon rank-sum test. The raw *P-*values were adjusted to the false discovery rate (FDR) using the Benjamini-Hochberg procedure. Gene transcripts with log_2_ |fold change| ≥ 1 and FDR ≤ 0.05 were considered to have significant differential coverages in the Ptss regions (Supplemental Table S5).

### Sample clustering and gene function annotation

The principal component analysis (PCA) was performed using the rgl package (ver. 0.1). Hierarchical clustering of the coverage data with the complete linkage clustering algorithms was implemented using the pheatmap package (ver. 1.0.2). The enrichment analysis of Gene Ontology (GO) and KEGG was completed using Metascape (ver. 20220101) ^25^ and clusterProfiler (ver. 3.18.1) ^26^ with their default settings. To construct the gene correlation network, the relationship of gene function was obtained from the string database (ver. 11.5) ^27^ and then the network was drawn with Cytoscape (ver. 3.8). To find the hub genes in the network, the gene degree of the network was analyzed using Cytohubba (ver. 0.1) ^28^.

### Predictive classifier construction and validation

To develop classifiers for predicting preterm birth (Figure 4), we implemented whole-genome sequencing of cfDNA on 2,590 pregnant women, including 518 preterm and 2,072 full-term pregnancies from three independent hospitals, including JM, FS, and NFY. The samples collected from JM (n=1,310) were randomly divided into training (n=915, training cohort [n=915]: 183 cases and 732 controls) and internal validation cohorts (validation1 cohort [n=395]: 79 cases and 316 controls) at a ratio of 7:3, while the samples collected from FS (validation2 cohort [n=930]: 186 cases and 744 controls) and NFY (validation3 cohort [n=350]: 70 cases and 280 controls) were taken as external validation cohorts (Figure 4). The clinical characteristics of the preterm and full-term pregnancies were well matched among four cohorts (Table 1). Since many studies have reported that discrete data may improve the predictive performance ^29^, before the classifiers were built, the read coverage of each pTSS identified in the discovery cohort was discretized according to the optimal cut-off point with the largest AUC value in the training cohort (Supplemental Table S7). The read coverage for each promoter in each subject was then set to one when it was larger than the corresponding optimal cut-off; Otherwise, it was set to zero. Then the sigFeature package (ver. 1.8.0) was used to evaluate the importance of the pTSS regions.

We then evaluated three predictive models, including support vector machine (SVM), logistic regression (LR), and linear discriminant analysis (LDA) as the base for developing a novel predictive classifier for PTB. The SVM model was constructed using the linear kernel in the e1071 package (ver. 1.7.9) with the default settings. While the glm and lda functions from the MASS package (ver. 7.3.53) were used to develop LR and LDA classifiers. We then combined each of these predictive models with either backward or lasso algorithms for feature selection. For lasso feature selection, the best lamda was identified using 10-fold cross validation with the cv.glmnet of glmnet package (ver. 4.1) and then the features with non-zero coefficients were selected. Finally, classifiers for preterm birth prediction were developed. The robustness of trained classifiers was assessed using the leave-one-out cross validation method (LOOCV). Briefly, each subject in the training cohort was withheld in turn, and the remaining subjects were submitted to the training classifier. The trained classifier was used to predict the class of the withheld subject. This process continued until all subjects in the training cohort have been judged. According to the AUC after cross-validation (CV), the classifier with the largest AUC, named PTerm, in the training cohort was selected.

To further assess the performance of PTerm, the whole-genome sequencing data from one internal validation (JM, validation1 cohort) and two external validation cohorts (FS, validation2 cohort; NFY, validation3 cohort), was implemented. The performance of PTerm was further evaluated using the data from these internal and external cohorts.

### Statistical analyses

Wilcoxon rank-sum test was used to compare the continuous variables between the preterm and full-term groups, while Pearson’s χ^2^ and Fisher’s exact tests were used for comparisons of categorical variables. The Wilcoxon rank-sum test was used to identify genes with differential read coverages within the pTSS regions and *P*-values of < 0.05 in two-sided tests were considered to be statistically significant. ROC curves and the significant differences in their AUC, sensitivity and specificity were plotted and calculated using the pROC package in R.

## Data Availability

All data produced in the present study are available upon reasonable request to the authors

## Acknowledgements

We thank Sagene eBioart for their help in making the pattern diagram.

## Author contributions

J T, X-x Y, F Y, Y-s W and X H designed and supervised the study. Z-w G, K W and K L analyzed and interpreted the data and prepared the manuscript. H-h S, J-y T and L-p L designed the study, provided samples and interpreted clinical data. X Y, M Z, B-w H and X-m Z analyzed and interpreted the data. X-c Z provided suggestions for manuscript revision. All authors vouched for the respective data and analysis, reviewed and approved the final version and agreed to publish the manuscript.

## Reference

1. Chawanpaiboon S, et al. Global, regional, and national estimates of levels of preterm birth in 2014: a systematic review and modelling analysis. Lancet Glob Health 7, e37–e46 (2019).

2. Crump C, Sundquist J, Sundquist K. Preterm delivery and long term mortality in women: national cohort and co-sibling study. BMJ 370, m2533 (2020).

3. Liang L, et al. Metabolic Dynamics and Prediction of Gestational Age and Time to Delivery in Pregnant Women. Cell 181, 1680–1692 e1615 (2020).

4. Ngo TTM, et al. Noninvasive blood tests for fetal development predict gestational age and preterm delivery. Science 360, 1133–1136 (2018).

5. Tarca AL, et al. Crowdsourcing assessment of maternal blood multi-omics for predicting gestational age and preterm birth. Cell Rep Med 2, 100323 (2021).

6. Guo ZW, et al. Noninvasive prediction of response to cancer therapy using promoter profiling of circulating cell-free DNA. Clin Transl Med 10, e174 (2020).

7. Wong FC, Lo YM. Prenatal Diagnosis Innovation: Genome Sequencing of Maternal Plasma. Annu Rev Med 67, 419–432 (2016).

8. Romero R, Dey SK, Fisher SJ. Preterm labor: one syndrome, many causes. Science 345, 760–765 (2014).

9. Leung TN, Zhang J, Lau TK, Chan LY, Lo YM. Increased maternal plasma fetal DNA concentrations in women who eventually develop preeclampsia. Clin Chem 47, 137–139 (2001).

10. Snyder MW, Kircher M, Hill AJ, Daza RM, Shendure J. Cell-free DNA Comprises an In Vivo Nucleosome Footprint that Informs Its Tissues-Of-Origin. Cell 164, 57–68 (2016).

11. Ulz P, et al. Inferring expressed genes by whole-genome sequencing of plasma DNA. Nat Genet 48, 1273–1278 (2016).

12. Esfahani MS, et al. Inferring gene expression from cell-free DNA fragmentation profiles. Nat Biotechnol 40, 585–597 (2022).

13. Guo Z, et al. Whole-Genome Promoter Profiling of Plasma DNA Exhibits Diagnostic Value for Placenta-Origin Pregnancy Complications. Adv Sci (Weinh) 7, 1901819 (2020).

14. Rubens CE, Sadovsky Y, Muglia L, Gravett MG, Lackritz E, Gravett C. Prevention of preterm birth: harnessing science to address the global epidemic. Sci Transl Med 6, 262sr265 (2014).

15. Flenady V, Reinebrant HE, Liley HG, Tambimuttu EG, Papatsonis DN. Oxytocin receptor antagonists for inhibiting preterm labour. Cochrane Database Syst Rev, CD004452 (2014).

16. Fernando F, et al. The idiopathic preterm delivery methylation profile in umbilical cord blood DNA. BMC Genomics 16, 736 (2015).

17. Zhang G, et al. Genetic Associations with Gestational Duration and Spontaneous Preterm Birth. N Engl J Med 377, 1156–1167 (2017).

18. Kaitu’u-Lino TJ, et al. Activating Transcription Factor 3 Is Reduced in Preeclamptic Placentas and Negatively Regulates sFlt-1 (Soluble fms-Like Tyrosine Kinase 1), Soluble Endoglin, and Proinflammatory Cytokines in Placenta. Hypertension 70, 1014–1024 (2017).

19. Rasmussen M, et al. RNA profiles reveal signatures of future health and disease in pregnancy. Nature 601, 422–427 (2022).

20. Moufarrej MN, et al. Early prediction of preeclampsia in pregnancy with cell-free RNA. Nature 602, 689–694 (2022).

21. Samura O. Update on noninvasive prenatal testing: A review based on current worldwide research. J Obstet Gynaecol Res 46, 1246–1254 (2020).

22. Scharfe-Nugent A, et al. TLR9 provokes inflammation in response to fetal DNA: mechanism for fetal loss in preterm birth and preeclampsia. J Immunol 188, 5706–5712 (2012).

23. Casper J, et al. The UCSC Genome Browser database: 2018 update. Nucleic Acids Res 46, D762–D769 (2018).

24. Turturice BA, et al. Perinatal granulopoiesis and risk of pediatric asthma. Elife 10, (2021).

25. Zhou Y, et al. Metascape provides a biologist-oriented resource for the analysis of systems-level datasets. Nat Commun 10, 1523 (2019).

26. Wu T, et al. clusterProfiler 4.0: A universal enrichment tool for interpreting omics data. Innovation (N Y) 2, 100141 (2021).

27. Szklarczyk D, et al. The STRING database in 2021: customizable protein-protein networks, and functional characterization of user-uploaded gene/measurement sets. Nucleic Acids Res 49, D605–D612 (2021).

28. Otasek D, Morris JH, Boucas J, Pico AR, Demchak B. Cytoscape Automation: empowering workflow-based network analysis. Genome Biol 20, 185 (2019).

29. Lin XJ, et al. A serum microRNA classifier for early detection of hepatocellular carcinoma: a multicentre, retrospective, longitudinal biomarker identification study with a nested case-control study. Lancet Oncol 16, 804–815 (2015).

