## Supplemental Materials for "Whole-Genome Promoter Profiling of Plasma Cell-Free DNA Exhibits Predictive Value for Preterm Birth"

**Supplemental methods**

**The definition of preterm birth (PTB) and full-term pregnancies**

The plasma samples of pregnancies were collected at 12–28 weeks of gestation. These samples were then retrospectively assigned to a birth outcome group based on their subsequent delivery time with PTB defined as birth < 37 weeks of gestation. In this study, the gestation age of full-term controls in our study was ≥ 38 weeks of gestation. For full-term pregnancies, the pregnancies with pregnancy complications were excluded. In addition, all participants were singleton pregnancies and pregnancies were excluded pregnancies were excluded: (1) chromosomal or congenital abnormalities; (2) infection; (3) pregnancies with uterine fibroids, or uterine malformation; (4) history of heparin, aspirin, or other drug use. Gestational age was determined according to the last menstrual period.

**Sample collection and cfDNA isolation**

Maternal whole blood was collected using Streck cell-free DNA blood collection tubes (BCT; Streck, USA) and was then centrifuged at 1,600g and 25°C for 15 minutes before plasma collection. This plasma was then centrifuged a second time at 2,500 g and 25°C for 10 minutes. After the second spin, the plasma was removed from the pellet that formed at the bottom of the tube and distributed into 4 mL barcoded plasma aliquots and immediately stored frozen at −80°C until DNA extraction. Then, according to the manufacturer’s instructions, cfDNA was extracted from plasma samples by using the QIAamp DNA blood mini kit (Qiagen, Germany). Qubit fluorometer (ThermoFisher Scientific, USA) and Agilent 2100 bioanalyzer (Agilent Technologies, USA) were used to measure DNA concentration and integration. The DNA was eluted in 50 µL AE buffer and stored at −20°C.

**Whole-genome Sequencing of plasma cfDNA**

Plasma cfDNA libraries were prepared following the Illumina TruSeq library preparation protocol (Illumina, USA) and AMPure XP magnetic bead clean-up (Beckman Coulter, USA) on a Caliper Zephyr liquid handler (PerkinElmer, USA). The TruSeq indexes were then incorporated into the libraries before each was quantified on the Caliper LabChip GX (PerkinElmer, USA) and normalized to the same concentration These libraries were sequenced using the NextSeq platform (Illumina, USA). For each sample, approximately four million 36-bp reads were generated.

**The selection of full-term controls in this case-control study**

This is a nested case-control study. According to other case-control studies published in journals with high impact factor,^1-3^ the ratio of control and case is usually 3–5:1. Based on the number of gestation age-matched full-term controls in our cohort, we selected four full-term controls for each preterm pregnant woman. The clinical characteristics of preterm birth and their corresponding controls matched well in four cohorts (Table 1; all *P*-value > 0.05, Mann-Whiney U test).

**Reference**

1.	Grimes DA, Schulz KF. Compared to what? Finding controls for case-control studies. *Lancet* 2005; **365**(9468): 1429-33.

2.	Hernan MA, Wilcox AJ. Epidemiology, data sharing, and the challenge of scientific replication. *Epidemiology* 2009; **20**(2): 167-8.

3.	Lu Y, Lagergren J, Eloranta S, Lambe M. Childbearing and salivary gland cancer: a population-based nested case-control study. *Epidemiology* 2009; **20**(5): 780-2.

**A list of Supplemental Figures and Supplemental Tables**

| ID | Description | File |
| --- | --- | --- |
| Supplemental Figure 1 | cfDNA profiles at promoter region reflect nucleosome positioning of blood cells in pregnancies | In this file |
| Supplemental Table S1 | 500 highest/lowest expressed genes in placenta | In a separated excel file |
| Supplemental Table S2 | Placenta- and whole blood-specific genes | In a separated excel file |
| Supplemental Table S3 | Housekeeping and unexpressed genes | In a separated excel file |
| Supplemental  Table S4 | Clinical characteristics of pregnancies in the discovery cohort | In this file |
| Supplemental Table S5 | Gene transcripts with differential read coverages at the pTSS | In a separated excel file |
| Supplemental Table S6 | Functional annotation of hub genes by retrieving literature | In this file |
| Supplemental Table S7 | The cut-off of gene promoter coverages for discretion | In a separated excel file |
| Supplemental Table S8 | Performance of the optimal classifier for each model with backward and lasso algorithm | In this file |
| Supplemental Table S9 | Functional annotation of genes in PTerm by retrieving literatures | In this file |

**Supplemental Figures and legends**

**Supplemental Figures**

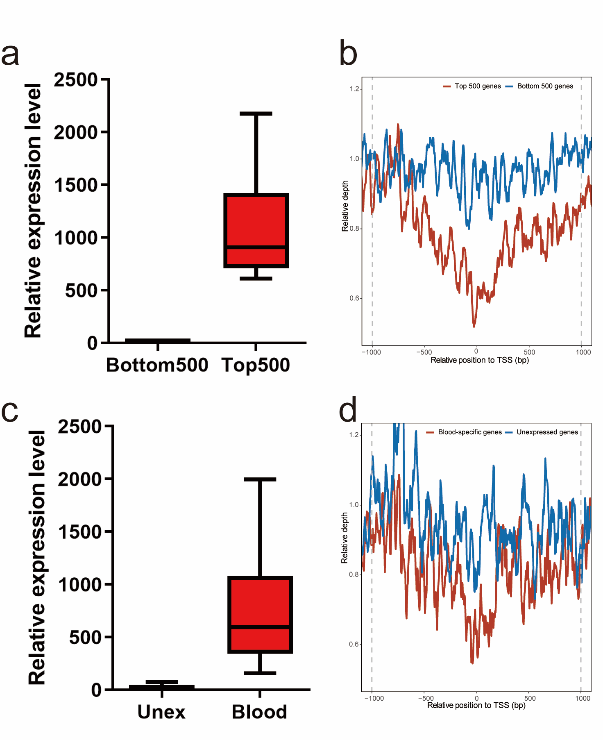

**Supplemental Figure 1. cfDNA profiles at promoter region reflect nucleosome positioning of blood cells in pregnancies. (a)** Average expression levels of the most highly expressed genes (Top500, red) and the 500 least expressed genes (Bottom500, blue) in the blood of preterm birth pregnancies. **(b)** Read depth of whole-genome sequencing at the pTSS region (-1KB to 1KB around the TSS) of the 500 most highly expressed genes (Top500, red line) and the 500 least expressed genes (Bottom500, blue line). **(c)** Average expression levels of the blood-specific genes (blood, red) and the unexpressed genes (Unex, blue) in the blood of preterm birth pregnancies. **(d)** Read depth of whole-genome sequencing at pTSS region (-1KB to 1KB around TSS) of the blood-specific genes (red line) was lower than that of unexpressed genes (blue line). The RNA expression profiling of whole blood of preterm birth was downloaded from GEO (GSE73685). The list of blood-specific genes, unexpressed genes, top500 and bottom500 genes were shown in Supplemental Table S1-S3. pTSS region (-1KB to 1KB around the TSS) was marked between grey dash lines. PTB, preterm birth; TSS, transcriptional start site; cfDNA, cell-free DNA.

**Supplemental Tables**

**Supplemental Table S4. Clinical characteristics of pregnancies in discovery cohort**

| Characteristics | Preterm (n=20) | Full-term (n=20) | *P-*value |
| --- | --- | --- | --- |
| Gestational age at sampling (weeks) | 15.7±4.3 | 15.3±3.0 | 0.386 |
| Maternal age (years) | 30.6±4.9 | 30.1±2.6 | 0.946 |
| BMI (kg/m^2^) | 21.0±2.5 | 20.9±3.6 | 0.626 |

Data are mean ± standard deviation. Age = maternal age. BMI = pre-pregnancy body mass index. Wilcoxon rank-sum test was used for the comparison of continuous variables.

**Supplemental Table S6. Functional annotation of the hub genes by retrieving literature**

| Gene | Function annotation | PMID |
| --- | --- | --- |
| *ERBB2* | 1. The catalytic activity of *ErbB-2* is essential for normal embryonic development. 2. *ERBB2* gene amplification increases during the transition of proximal EGFR+ to distal HLA-G+ first trimester cell column trophoblasts. | 1. 11809799 2. 26071215 |
| *ESR1* | 1. *ESR1* gene polymorphism is associated with preterm rupture of fetal membranes. 2. The methylation trend of the majority of gestational age related DMPs increases with advancing gestation and within this set are 3 DMPs localising to the *ESR1* gene. 3. Estrogen production may play a role in the stimulation of cytotrophoblast cell differentiation and promote function in normal placentae. | 1. 24824712 2. 26419829 3. 12646062 |
| *NFKBIA* | 1. The expression levels of inflammation-inhibiting gene, *NFKBIA*, was up-regulated in the preterm pregnancies | ① 35199834 |
| *HSPA5* | 1. The WES datasets also supported roles for HSP and NR genes in SPTB susceptibility | ① 34429451 |
| *PRKCB* | 1. Pregnancy is associated with a marked upregulation in the expression of native and activated forms of the *PRKCB* dependent inhibitory phospho-protein CPI-17. | ① 19589855 |
| *RAF1* | 1. A Novel Noonan Syndrome *RAF1* Mutation: Lethal Course in a Preterm Infant 2. *RAF1* expression is increased in myometrium with human labour and mediators of preterm labour, and inhibition of *RAF1* | 1. 26266034 2. 26811545 |
| *NFE2L2* | 1. Association between SNPs in *NFE2L2* involved in folate metabolism and preterm birth risk | ① 25730024 |
| *SNAI1* | 1. *SNAI1* regulate embryonic development and involved in trophoblast differentiation. | 1. 22360878 |
| *GSN*  (*Gelsolin*) | 1. Low plasma gelsolin levels in the first postnatal month may be associated with poor outcomes in premature infants. | ① 24862494 |
| *ATF3* | 1. *ATF3* is downregulated in preterm preeclampsia and is negatively related to sflt1 2. The expression of *ATF3* is significantly decreased in preterm placentas | ① 28947613  ② 26880734 |

PMID = PubMed Unique Identifier.

**Supplemental Table S8. Performance of the optimal classifier for each model with backward and lasso algorithm**

|  | Backward | Lasso | *P*-value |
| --- | --- | --- | --- |
| SVM | | | |
| AUC (95% CI) | 0.88 (0.85-0.90) | 0.73 (0.69-0.77) | 2.2e-16 |
| LDA | | | |
| AUC (95% CI) | 0.80 (0.76-0.83) | 0.74 (0.70-0.78) | 2.2e-16 |
| LR | | | |
| AUC (95% CI) | 0.86 (0.83-0.88) | 0.69 (0.66-0.73) | 2.2e-16 |

The significant differences in their AUC were compared using DeLong's test. SVM=support vector machine; LR=logistic regression; LDA=linear discriminant analysis.

**Supplemental Table S9. Functional annotation of genes in PTerm by retrieving literatures**

| Gene | Function annotation | PMID |
| --- | --- | --- |
| *ERBB2* | 1. The catalytic activity of *ErbB-2* is essential for normal embryonic development. 2. *ERBB2* gene amplification increases during the transition of proximal EGFR+ to distal HLA-G+ first trimester cell column trophoblasts. | 1. 11809799 2. 26071215 |
| *NFKBIA* | ①The expression levels of inflammation-inhibiting gene, NFKBIA, was up-regulated in the preterm pregnancies | ① 35199834 |
| *RAF1* | 1. A Novel Noonan Syndrome *RAF1* Mutation: Lethal Course in a Preterm Infant 2. *RAF1* expression is increased in myometrium with human labour and mediators of preterm labour, and inhibition of *RAF1* | 1. 26266034 2. 26811545 |
| *GSN*  (*Gelsolin*) | 1. Low plasma gelsolin levels in the first postnatal month may be associated with poor outcomes in premature infants. | ① 24862494 |
| *TMEM9B* | Regulation of differentiation of Bone Marrow-Derived Mesenchymal Stem Cells | 1. 33299881 |
| *TARBP2* | 1. Significantly expressed in astrocyte maturation | ① 14703617 |
| *ZIC2* | 1. Aberrant signaling involving the *ZIC2* and *TGIF* genes are common causes of human Holoprosencephaly | ① 24753843 |
| *CYP1B1* | 1. *CYP1B1* is significantly more expressed in preterm labor patients | 1. 23916819 |
| *RPS6KB1* | 1. Deficiency of the oxidative stress–responsive kinase p70S6K1 restores autophagy and ameliorates neural tube defects in diabetic embryopathy | 1. 32416155 |
| *RIPK1* | 1. RIPK1 mRNA level was significantly increased in PE placentas. | 1. 28292463 |
| *FSCN1* | 1. miR-143 and miR-145 were significantly increased in cervical cells of women with PTB. miR-143 and miR-145 transfection decreased cervical cell number by increasing apoptosis and decreasing cell proliferation through initiation of cell cycle arrest. Cell adhesion genes, JAM-A and *FSCN1*, were downregulated with overexpression of miR-143 and miR-145. | 1. 28596604 |
| *RIT1* | 1. The mutations of *RIT1* were associated with Noonan syndrome | 1. 27109146 |
| *CHKA* | Polymorphic variants of *CHKA* involved in choline pathway and the risk of intrauterine fetal death | ① 28509322 |
| *NFATC4* | 1. *NFATC4* is required for cardiac development and mitochondrial function | 1. 12750314 |
| *THRA* | 1. Termination of pregnancy with mifepristone leads to a downregulation of THRα1, THRα2 and THRβ1 in villous trophoblasts and in addition to a decreased expression of THRA in placental tissue. Decreased expression of THRα1 induced by RU486 could also be found in the decidua. | ① 26476797 |
| *MAP3K3* | 1. Appropriate activation of *MAP3K3* can play important roles in pregnancy | 1. 32175444 |
| *KCNJ2* | The mutations of *KCNJ2* results in Andersen Syndrome | ① 12148092 |
| *SNTA1* | 1. *SNTA1* is associated with long-QT syndrome | ① 18591664 |
| *TTC7A* | 1. *TTC7A* Mutation in a Newborn with Multiple Intestinal Atresia and Combined Immunodeficiency | 1. 25546680 |
| *PLXNB1* | 1. A decreased expression of PLXNB1 in preeclamptic placentas may be responsible for the deficiency in Met signaling and in PE development | ① 29939944 |
| *CDC25C* | 1. The activation of the ATR-CDC25C-CDK1 pathway induces cell cycle arrest at G2-phase. | ① 28264028 |
| *SKAP2* | 1. Placental epigenome-wide association study identified loci associated with childhood Adiposity. Four candidate epigenomic regions associated with skinfold thickness, which were located within *FMN1*, *MAGI2*, *SKAP2* and *BMPR1B* genes. | 1. 33003475 |
| *OGDH* | 1. A biallelic pathogenic variant in the *OGDH* gene results in a neurological disorder with features of a mitochondrial disease. | ① 32383294 |
| *SOX8* | 1. *SOX8* takes part in mammalian testis development | ① 19647095 |
| *PSMD3* | 1. *PSMD3* takes part in hedgehog ligand biogenesis, which is crucial for the development and differentiation | ① 34674377 |
| *PRKCQ* | ①*PRKCQ* involved in the development and differentiation of early innate lymphoid progenitors | ① 29183988 |
| *CDH22* | ①*Cdh12* and *Cdh22* in the developing and adult mouse brain | 1. 20723620 |
| *SPDYA* | 1. The *SUN1*-*SPDYA* interaction plays an essential role in meiosis prophase I in cell cycle. | ① 34039995 |

PMID = PubMed Unique Identifier.
